# Intra-Patient Comparison of a Minimally Invasive Lid Treatment and Traditional Blepharoplasty: Patient and Surgeon-Reported Outcomes

**DOI:** 10.64898/2026.10.02.26364300

**Authors:** Ana Carolina Victoria, Richard E. Roth, Mark N. Berkowitz, Sumeet Jindal, Patricia Buehler

## Abstract

**Purpose:** To use a minimally invasive lid treatment (MILT) utilizing Compressive Skin Contouring (CoCo) technology to compare patient- and surgeon-reported ecchymosis, edema, and incision appearance following MILT and traditional upper eyelid blepharoplasty (tBULB).

**Methods:** Prospective, single-surgeon intra-patient study comparing MILT and tBULB in patients seen for bilateral blepharoplasty. Patients and the operating surgeon rated MILT and tBULB outcomes post-operatively on the day of surgery (T1) and at 2 months (T2) using surveys. Patient photographs taken 3 and 9 days post-operatively were blindly assessed by 2 independent surgeons to compare ecchymosis, edema, and incision appearance on the MILT and tBULB sides.

**Results:** Seven patients participated in the study. At T1, 6/7 patients reported ≤ ecchymosis and edema, and all patients reported better or equivalent incision appearance on the MILT side compared to the tBULB side. At T2, 6/7 of patients reported ≤ edema; 4/7 reported < ecchymosis and better incision appearance on the MILT side. The surgeon reported < ecchymosis and ≤ edema for 7/7 patients and better or equivalent incision appearance for 6/7 patients at T1; ≤ ecchymosis and better or equivalent incision appearance for 7/7 and ≤ edema for 6/7 patients at T2 on the MILT side. In the blinded evaluation, mean ecchymosis scores were lower, edema scores were lower or equivalent, and incision appearance was better for MILT vs. tBULB at days 3 and 9.

**Conclusions:** In this intra-patient study, MILT outcomes compared favorably with tBULB, especially for ecchymosis and edema; two important aesthetic measures of patient perception.

## Introduction

The eye is a key component of facial aesthetics, and blepharoplasty can play a vital positive role in patients’ perception of aging. Some common changes that occur with aging are loss of volume in the upper eyelid and periorbital areas, a higher position of the eyelid crease, a lower position of the eyebrow, and dermatochalasis.^1^ Patients with these symptoms may benefit from upper eyelid blepharoplasty to revitalize facial appearance. Upper eyelid blepharoplasty is one of the most performed facial cosmetic procedures in the United States, with more than 320,000 procedures performed in 2020.^2^ Although side effects following traditional upper eyelid blepharoplasty (tBULB) generally resolve within a few months, patients may experience ecchymosis, edema, and visible scarring for several weeks.^3^ Fear of harms due to the blepharoplasty procedure (cutting, cautery, suturing, and suture removal), surgical outcomes (asymmetry and wound dehiscence), as well as post-operative ecchymosis and edema concerns may influence an individual’s decision to undergo tBULB.

To mitigate some of the potential patient harms associated with tBULB, we developed a Minimally Invasive Lid Treatment (MILT) that utilizes proprietary Compressive Skin Contouring (CoCo) technology (Ziplyft^®^, Osheru, Inc., Bend, OR) for upper eyelid blepharoplasty. CoCo technology utilizes skin compression to cut off blood supply locally, thus eliminating the need for cautery and bleeding management during the blepharoplasty procedure while also reducing the potential for ecchymosis. CoCo technology also seals the skin edges together taking tension off the wound to allow for closure with commercially available tissue adhesive instead of sutures. The surgical improvements associated with MILT have the potential to make the blepharoplasty procedure significantly less invasive for the patient. The anticipated benefits include reduced blepharoplasty procedure complications, especially those related to wound dehiscence and scaring, and reduced postoperative limitations due to prolonged ecchymosis and edema. In this prospective intra-patient split face pilot study, we compared patient-and surgeon-reported measures of ecchymosis, edema, and incision appearance following MILT and tBULB. Clinical Trial Identifier: NCT07250139

## Methods

### Study Design and Participants

This prospective, single-surgeon prospective intra-patient comparative study enrolled male and female patients > 35 years of age being seen for bilateral upper eyelid blepharoplasty at a participating eyecare center. Patients received MILT on one side and tBULB on the contralateral side of their face. Patients were randomized with respect to which side of the face the MILT or tBULB procedure was performed on. The study and data collection was conducted with Institutional Review Board approval, and the study was conducted in accordance with Declaration of Helsinki and Health

Insurance Portability and Accountability Act (HIPPA) regulations. All eligible patients provided written informed consent prior to study participation. All patients who participated in the study provided written consent for the use of facial photographs for comparative assessments and for publication; these signed consent forms are on file at the participating surgeon’s office. Patients who could not provide informed consent to participate in the study were excluded. Additionally, patients with Ptosis or Myasthenia Gravis, who were pregnant or nursing, had a history of Graves’ Disease, allergies to tissue adhesives, were active or recent (within 3 months) tobacco users, used anticoagulation therapy within 3 days prior to the study procedure, had any previous eyelid surgery or eyebrow region, participated in any study of an investigational, interventional product within 30 days prior to the preoperative visit or at any time during the study period, and those who, in the opinion of the investigator, were anatomically not conducive to using devices (e.g., heavy brow, severe hollowing of the upper eyelid, history of ocular or orbital trauma) were excluded from participating in the study.

### MILT Device Design and Surgical Procedure

The MILT device and procedure are shown in Figure 1 using simulated images. Oral valium is administered for conscious sedation and muscle relaxation. The unwanted excess skin to be removed from each lid is marked on both right and left sides. The clear plastic design of the MILT device allows the surgeon to see the skin markings and confirm the amount of tissue to be removed. The device has markings to ensure symmetric placement between the two eyelids. The MILT device with CoCo technology compresses the marked excess skin in the jaws of the clamp prior to removal, reducing bleeding and bruising. A guarded blade excises the excess, marked skin and the device is removed. The residual zipper-locked ridge of skin is tension free and can be sealed together using commercially available tissue adhesive, eliminating the need for suturing and suture removal.

**Figure 1.**
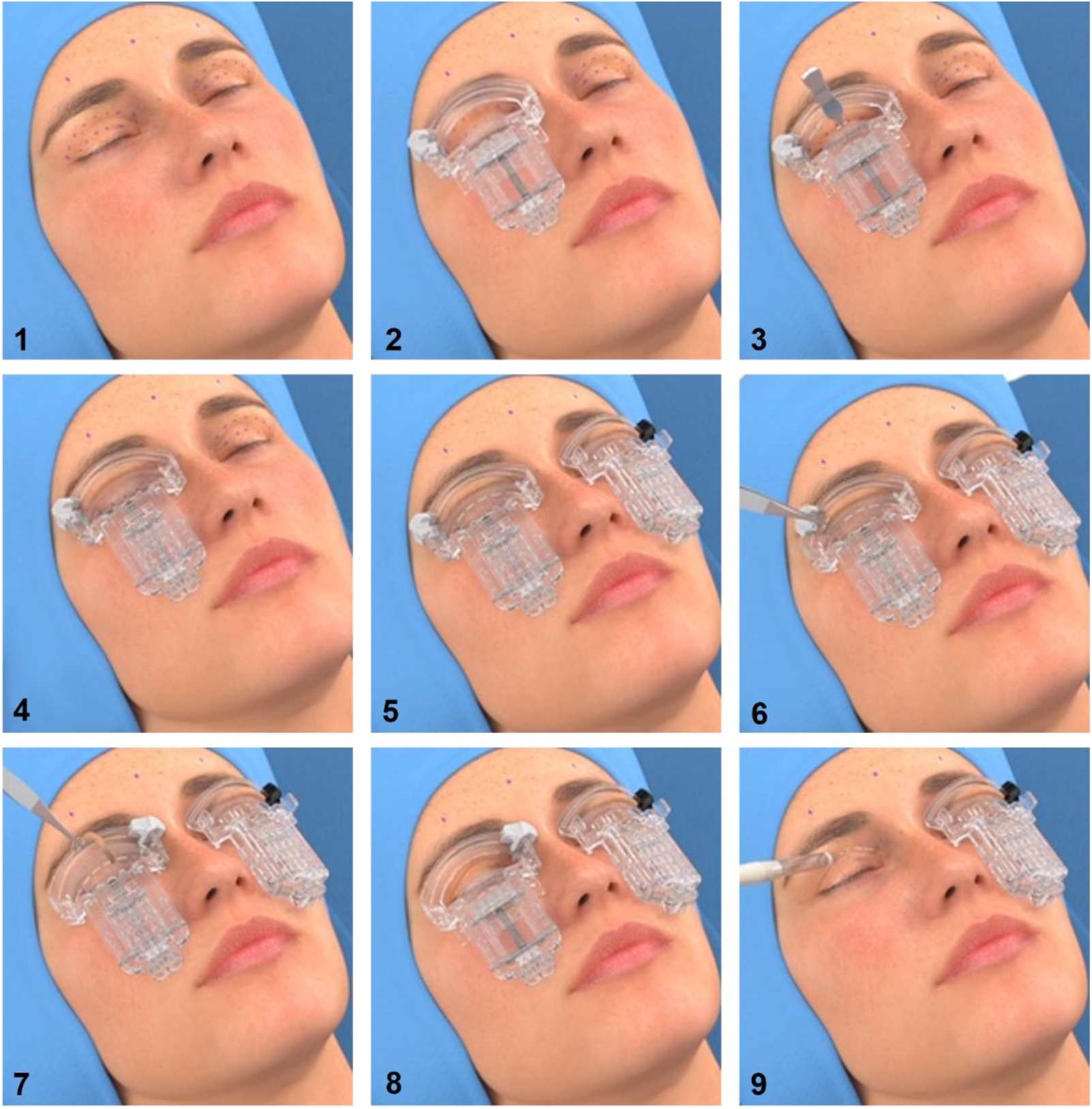
MILT device and procedure steps using simulated images. MILT, Minimally invasive lid treatment with compressive skin contouring (CoCo) technology. 1. Before the MILT procedure, the excess skin is marked for removal. 2. The MILT device is transparent, so skin markings are visible. It is contoured to align with the natural eyelid crease. 3. Excess skin is engaged using forceps and pulled into the compression jaw. 4. The compression jaw closes off small blood vessels, significantly reducing bleeding and eliminating the need for cauterization. 5. The right MILT device has a white blade handle, and the left device has a black blade handle. 6-7. Once compressed, the integrated guarded blade slides smoothly across the treatment area. 8. Once the skin removal is complete, the latch is released leaving a sealed ridge of tissue. 9. A thin layer of tissue adhesive is applied to achieve a tension-free closure.

### Traditional Upper Eyelid Blepharoplasty Procedure

A traditional skin-only blepharoplasty procedure was performed on the contralateral upper eyelid, utilizing a scalpel to remove the skin, cautery to eliminate bleeding, and sutures to close the incision.^3^

### Patient and Surgeon Reported Outcomes

Patients and the operating surgeon completed surveys post-operatively on the day of surgery and at two months post-operatively, comparing assessments of ecchymosis, edema, and incision appearance for the MILT and tBULB sides of the face.

### Independent Blinded Outcome Assessment

Patient photographs were taken on 3 and 9 days post-operatively were blindly assessed by two independent oculoplastic surgeons to compare ecchymosis, edema, and incision appearance on the MILT and tBULB sides of the face. Photographs were taken under identical lighting conditions, with the patient sitting upright looking straight into the camera with eyes open and with eyes closed. The surgeons used the Surgeon Periorbital Rating of Edema and Ecchymosis (SPREE) questionnaire to objectively score ecchymosis and edema.^4^ The SPREE questionnaire has been previously used as a grading scale in blepharoplasty studies.^5,6^ This scoring system rates the degree of edema by giving a score according to coverage of iris by the swollen lid (grade 1–4) and rates the degree of lid ecchymosis according to its extent, whether on the inner third only, the inner 2/3, or the whole lid (grade 0–3). Incision appearance was scored using the Scar Cosmesis Assessment Rating (SCAR) scale, which includes measures (0-3 scale) of spread, erythema, dyspigmentation, suture marks, and hypertrophy.^7^ The surgeons graded randomized images without treatment assignment; ecchymosis and edema scores were averaged for each eye at each timepoint.

### Statistics

Comparative observations (MILT better, equivalent, or tBULB better) are reported for the patient and surgeon reported measures. Paired MILT and tBULB SPREE score comparisons were analyzed using the Wilcoxon signed-rank test.

## Results

### Study Participants

Seven patients (six female; one male) non-Hispanic, White patients participated in the study. Mean age was 50.0 years. All patients were being seen for aesthetic purposes.

### Patient Reported Outcomes

Patient reported assessments of their outcomes following MILT and tBULB are shown in Table 1. On the day of surgery, 6/7 patients reported less or equivalent ecchymosis and edema, and all patients reported better or equivalent incision appearance on the MILT side compared to the tBULB side. At two months post-op, 6/7 of patients reported less or equivalent edema; 4/7 reported less ecchymosis and better incision appearance on the MILT side compared to the tBULB side.

**Table 1.** Patient reported measures of ecchymosis, edema, and incision appearance.

| Assessment | Timepoint | MILT better | Equal | tBULB better |
| --- | --- | --- | --- | --- |
| Less ecchymosis | Day of Surgery | 57% (4/7) | 29% (2/7) | 14% (1/7) |
| Less ecchymosis | 2 Month | 57% (4/7) | 0% (0/7) | 43% (3/7) |
| Less edema / swelling | Day of Surgery | 71% (5/7) | 14% (1/7) | 14% (1/7) |
| Less edema / swelling | 2 Month | 57% (4/7) | 29% (2/7) | 14% (1/7) |
| Incision appearance | Day of Surgery | 29% (2/7) | 71% (5/7) | 0% (0/7) |
| Incision appearance | 2 Month | 57% (4/7) | 0% (0/7) | 43% (3/7) |

MILT, Minimally invasive lid treatment; tBULB, traditional upper eyelid blepharoplasty.

### Surgeon Reported Outcomes

Surgeon reported assessments of patient outcome following MILT and tBULB are shown in Table 2. The operating surgeon reported less ecchymosis and less or equivalent edema for 7/7 patients and better or equivalent incision appearance for 6/7 patients at T1; ≤ ecchymosis and better or equivalent incision appearance for 7/7 and ≤ edema for 6/7 patients at T2 on the MILT side.

**Table 2.** Surgeon reported measures of ecchymosis, edema, and incision appearance.

| **Assessment** | **Timepoint** | **MILT better** | **Equal** | **tBULB better** |
| --- | --- | --- | --- | --- |
| Less ecchymosis | Day of Surgery | 100% (7/7) | 0% (0/7) | 0% (0/7) |
| Less ecchymosis | 2 Month | 86% (6/7) | 14% (1/7) | 0% (0/7) |
| Less edema / swelling | Day of Surgery | 71% (5/7) | 29% (2/7) | 0% (0/7) |
| Less edema / swelling | 2 Month | 71% (5/7) | 14% (1/7) | 14% (1/7) |
| Incision appearance | Day of Surgery | 57% (4/7) | 29% (2/7) | 14% (1/7) |
| Incision appearance | 2 Month | 71% (5/7) | 29% (2/7) | 0% (0/7) |

MILT, Minimally invasive lid treatment; tBULB, traditional upper eyelid blepharoplasty.

### Independent Comparisons of MILT and tBULB Outcome

Days three and nine post-operative MILT vs. tBULB comparisons of ecchymosis, edema, and incision appearance/scaring are shown in Table 3. Average day three MILT ecchymosis scores were 0.93 points lower and day nine scores were 0.64 points lower than tBULB scores. Average day three MILT edema scores were 0.15 points lower and day nine scores were 0.21 points lower than tBULB scores. Average day three MILT incision appearance/scaring scores were 0.28 points lower and day nine scores were 0.17 points lower than tBULB scores. While MILT ecchymosis, edema, and incision appearance/scaring scores tended to be lower than tBULB scores, none reached significance. This is likely due to the small study sample size.

**Table 3.** Independent comparison of ecchymosis, edema, and incision appearance.

| Assessment | Timepoint | MILT, mean (±SD) | tBULB, mean (±SD) | Difference* | p-value |
| --- | --- | --- | --- | --- | --- |
| Ecchymosis | Day 3 | 1.21 (1.25) | 2.14 (1.03) | -0.93 | 0.297 |
| Ecchymosis | Day 9 | 0.43 (0.79) | 1.07 (1.10) | -0.64 | 0.406 |
| Edema | Day 3 | 1.64 (0.48) | 1.79 (0.49) | -0.15 | 0.750 |
| Edema | Day 9 | 1.36 (0.48) | 1.57 (0.53) | -0.21 | 0.562 |
| Incision Appearance/Scaring | Day 3 | 1.24 | 1.52 | -0.28 | 0.656 |
| Incision Appearance/Scaring | Day 9 | 1.67 | 1.83 | -0.17 | 0.953 |

\*Difference equals MILT average score minus tBULB average score. MILT, Minimally invasive lid treatment; tBULB, traditional upper eyelid blepharoplasty. p-values represent Wilcoxon signed-rank tests on paired within-subject differences.

## Discussion

This is the first report describing the procedure and outcomes following a MILT procedure for upper eyelid blepharoplasty using Compressive Skin Contouring (CoCo) technology. The tourniquet effect of the compression jaws of the MILT device, cuts off the blood supply to the skin, eliminates most bleeding and the need for cautery. Less bleeding, in addition to eliminating the multiple needle passes of suturing, has the potential to result in less bruising. Eliminating the inflammation of cautery and suturing may also contribute to less edema post operatively. This prospective, intra-patient pilot study was designed to compare ecchymosis, edema, and incision appearance following MILT and tBULB at short (one, three, and nine days post-op) and at longer-term, two-month post-operative follow-up. Since contralateral eyelids were operated on by the same oculoplastic surgeon during the same session, outcome measure comparisons between the two sides are attributable to the MILT or tBULB surgical procedures, rather than to patient characteristics, surgeon skill, or post-operative care.

Most patients reported less or equivalent ecchymosis and edema, and better or equivalent incision appearance for MILT compared to tBULB post-operatively on the day of surgery. These results agree with the surgeon-reported measures of ecchymosis, edema, and incision appearance for MILT on the day of surgery. Most patients also reported less or equivalent edema outcomes post-operatively at two-months follow-up for MILT, which is consistent with the surgeon’s assessment of edema at this timepoint. Patient and surgeon assessments differed with respect to ecchymosis and incision appearance measures at the two-month post-operative timepoint. Three of seven patients felt that ecchymosis and incision appearance were better on the tBULB side, while the surgeon reported better or equivalent incision appearance for all seven patients, and less or equivalent edema for six of seven patients on the MILT side. Several explanations for these divergent results are plausible. By two months post-operative follow-up, acute ecchymosis has likely resolved for both the MILT and tBULB sides of the face, so patient ratings of ecchymosis may reflect fine aesthetic distinctions that are highly individual. At two months post-op blepharoplasty scars are still in the softening and flattening phase.^8^ A surgeon assessment of scaring considers scar spread, erythema, dyspigmentation, suture marks, and hypertrophy, not just the cosmetic impression, which may influence the patient-reported outcomes. Even at three months post-op some degree of scar visibility has been reported in up to 23% of patients following tBULB.^9^. Independent surgeon assessments of day three and nine post-operative photographs consistently resulted in lower average ecchymosis scores, with slightly lower average scaring and slightly lower or equivalent average edema scores.

In addition to the patient-reported MILT outcomes noting better or equivalent ecchymosis and edema, and better or equivalent incision appearance for MILT compared to tBULB, surgeons also reported shortened case times with MILT. The average case time for the MILT procedure was 5 minutes and the average case time for the tBULB procedure was 7 minutes (data not shown). For a bilateral case, this equates to an average 10-minute bilateral MILT case time vs. an average 14-minute skin-only bilateral tBULB case time; nearly 30% reduction in case time for MILT vs. tBULB.

This study has some limitations. In this pilot study of seven patients, the small sample size likely contributed to the observed variations in patient- and surgeon-reported measures. The results reported herein should be viewed as qualitative, as the sample size prohibited rigorous statistical interrogation. The study was performed at a single site and the intra-patient MILT and tBULB procedures were performed by a single, experienced oculoplastic surgeon. While this scenario improves the validity of comparative measures, minimizing other variables besides the surgical techniques; broader multi-surgeon multi-site studies that include oculoplastic surgeons and general ophthalmologists would expand generalizability. The tBULB procedure was deliberately limited to skin-only removal with a scalpel; comparisons to other blepharoplasty variations may differ. Finally, future studies should include longer (6-12 months) follow-up to investigate the sustained benefit following MILT.

In summary, the present study provides preliminary patient- and surgeon-reported outcome measures comparing MILT and tBULB on the day of surgery and at two-months post-operatively. Most patients reported better or equivalent ecchymosis, edema, and incision appearance following MILT. Larger, multi-site MILT studies are ongoing.

## Data Availability

All data produced in the present study are available upon reasonable request to the corresponding author.

## Acknowledgements

The authors would like to thank Katie Gambino and the patients who participated in the study.

## Author Contributions

Conceptualization, Methodology, Formal Analysis, Resources, Formal Analysis, Validation, Project Administration: Patricia Buehler

Investigation, Data Curation: Ana Carolina Victoria, Richard E. Roth, Mark N. Berkowitz, Sumeet Jindal

Writing – original draft: Ana Carolina Victoria, Richard E. Roth, Mark N. Berkowitz, Sumeet Jindal, Patricia Buehler

Writing – review and editing: Ana Carolina Victoria, Richard E. Roth, Mark N. Berkowitz, Sumeet Jindal, Patricia Buehler

## References

1. Naik MN, Honavar SG, Das S, Desai S, Dhepe N. Blepharoplasty: an overview. J Cutan Aesthet Surg. Jan 2009;2(1):6–11. doi:10.4103/0974-2077.53092

2. American Society of Plastic Surgeons. Plastic Surgery Procedural Statistics Report. 2020. https://www.plasticsurgery.org/Documents/News/Statistics/2020/Plastic-Surgery-Statistics-Full-Report-2020.Pdf

3. Patel BC, R. M. Upper Eyelid Blepharoplasty. [Updated 2023 Jul 24]. StatPearls [Internet]. . StatPearls Publishing; 2026. https://www.ncbi.nlm.nih.gov/books/NBK537078/

4. Oliver JD, Menapace D, Younes A, Recker C, Hamilton G, Friedman O. Validation of the Modified Surgeon Periorbital Rating of Edema and Ecchymosis (SPREE) Questionnaire: A Prospective Analysis of Facial Plastic and Reconstructive Surgery Procedures. Facial Plast Surg. Feb 2018;34(1):95–101. doi:10.1055/s-0037-1615282

5. Marous CL, Farhat OJ, Cefalu M, Rothschild MI, Alapati S, Wladis EJ. Effects of Preoperative Intravenous Versus Subcutaneous Tranexamic Acid on Postoperative Periorbital Ecchymosis and Edema Following Upper Eyelid Blepharoplasty: A Prospective, Randomized, Double-Blinded, Placebo-Controlled, Comparative Study. Ophthalmic Plast Reconstr Surg. Sep-Oct 01 2024;40(5):523–532. doi:10.1097/iop.0000000000002633

6. Tharwat A, Saudi W, Solyman O, Elessawy K. Conventional vs noninvasive upper lid blepharoplasty in upper lid dermatochalasis: a comparative case series study. Delta Journal of Ophthalmology. 2020/07/01 2020;21(3):216–222. doi:10.4103/DJO.DJO_10_20

7. American Academy of Ophthalmology. EyeWiki. Scar Assessment Scales in Oculoplastic Surgery. Updated April 14, 2026. https://eyewiki.org/Scar_Assessment_Scales_in_Oculoplastic_Surgery

8. Aghajani A, Babaei L, Eshraghi B. From Incision to Healing: An Evidence-Based Review of Scar Optimization in Upper Blepharoplasty. Aesthetic Plastic Surgery. 2026/01/01 2026;50(2):604–615. doi:10.1007/s00266-025-05414-6

9. Vasović DD, Karamarković ML, Jovanović M, et al. Comprehensive Evaluation of Quality of Life following Upper Eyelid Blepharoplasty: A Prospective Analysis. Medicina (Kaunas). Mar 19 2024;60(3)doi:10.3390/medicina60030500

